# Individualized, parcel-guided rTMS to the left dorsolateral prefrontal cortex for treatment resistant depression

**DOI:** 10.64898/2026.09.03.26362155

**Authors:** Joshua Berman, Dennis Q. Truong, Andrew Murphy, Rashel Mejia, Yishai Z. Valter, Abhishek Datta, Daniel C. Javitt

## Abstract

Efficacy of Transcranial Magnetic Stimulation (TMS) for depression has been shown to correlate with the proximity of stimulation to the region of LDLPFC most anti-correlated with subgenual anterior cingulate cortex (sgACC). Using the Human Connectome Project (HCP) surface-based reconstruction from a structural MRI, we observed that the region of DLPFC most anti-correlated with sgACC falls in anterior parcel 46 and encompasses the junction between adjacent parcels. Individual parcel mapped TMS directed at this target showed high efficacy for treatment resistant depression in a group of 15 patients. Pre-treatment anti-correlation between DLPFC (parcel 46) and sgACC correlated significantly with treatment response. These findings, along with our prior study, demonstrate the feasibility of individual, parcel-guided rTMS and suggest response/remission rates similar to those observed using individualized fMRI localization of the peak anti-correlated region.

---

Repetitive transcranial magnetic stimulation (rTMS) is an effective therapy for treatment resistant depression with response and remission of rates 40-50% and 35%, respectively [1]. In clinical practice, stimulation is applied to the left dorsolateral prefrontal cortex (L-DLPFC) most commonly by targeting a scalp site either 5.5-cm anterior to motor cortex or at the F3 EEG electrode [2]. Treatment efficacy has been shown to correlate with the proximity of the stimulation site to the region of L-DLPFC that is most anti-correlated with subgenual anterior cingulate cortex (sgACC) [3-5]. However, optimal approaches for identifying this region prospectively in individual patients remain to be determined.

Here in an open label trial, we tested the utility of surface-based brain reconstruction and parcel-based targeting of rTMS using tools developed by the Human Connectome Project (HCP) including the Multimodal Parcellation (MMP) atlas [6]. We have previously observed that the peak sgACC anti-correlation region falls at the junction of MMP atlas parcels 46, 9-46d and a9-46v [7]. Moreover, open-label targeting of this region led to response and remission rates of 100% and 50% among individuals (n=10) non-responsive to conventional TMS [7].

For this study, we obtained data from 15 individuals with DSM-V moderate to severe major depressive disorder resistant to at least two adequate antidepressant trials, and with a current Montgomery-Asberg Depression Rating Scale (MADRS) rating score of ≥20 (**Fig. 1A**). The study was approved by Columbia University Medical Center (CUMC) and Western Institutional (WIRB) Review Boards and pre-registered online (NCT04956081). Patients were drawn from the CUMC psychiatry faculty practice and a departmental recruitment website. 12 of 15 (80%) patients were diagnosed with comorbid anxiety conditions. All participants received standard 10-Hz rTMS for 6 weeks (27 total sessions) using a CloudTMS Therapy System (Soterix Medical Inc, Woodbridge, NJ).

**Fig. 1.**
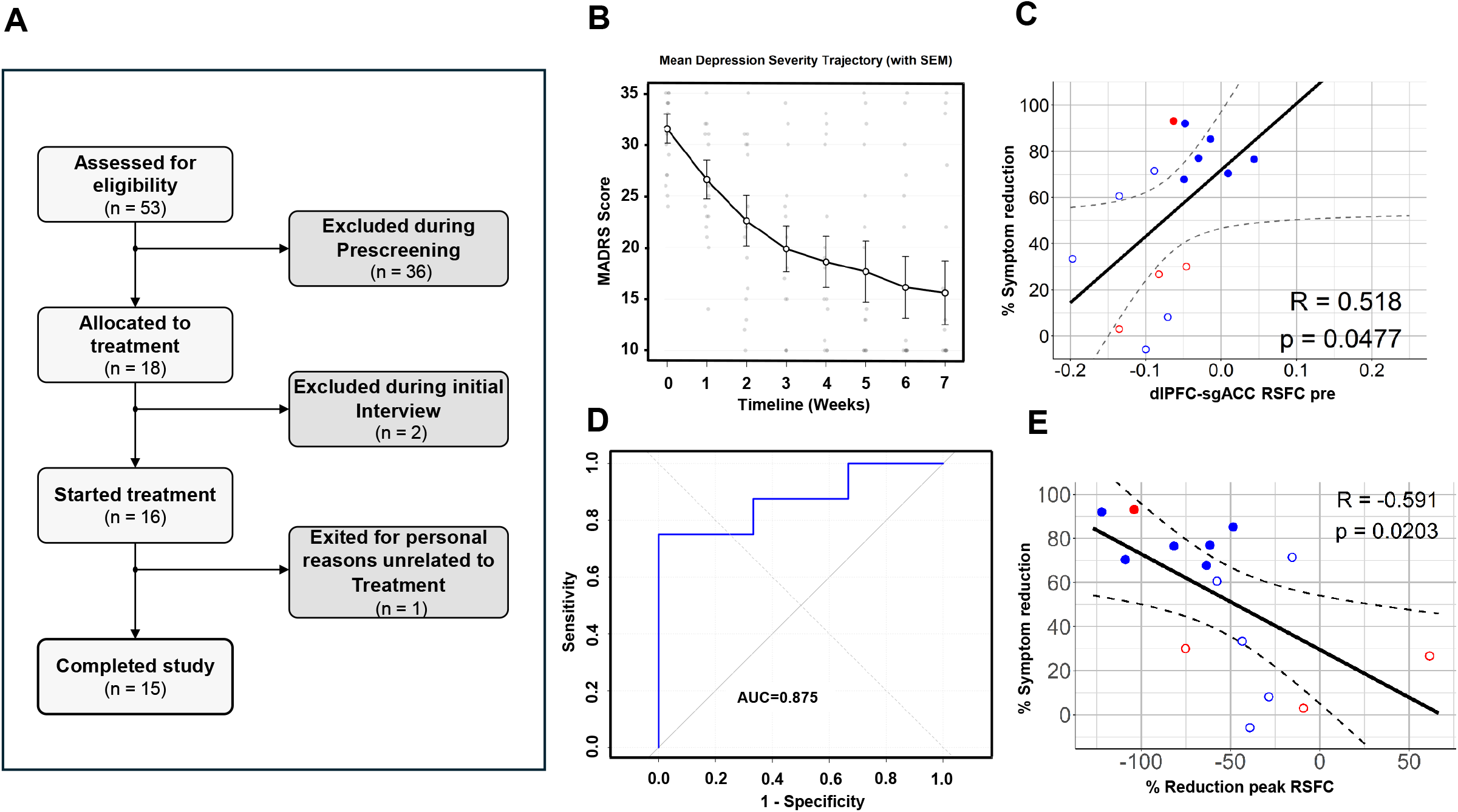
Parcel guided rTMS to the left DLPFC shows efficacy for treatment-resistant depression and connectivity between sgACC and LDLPFC Parcel 46 predicts response. **A)** 53 subjects assessed with 35 excluded, 18 allocated for treatment and 2 excluded during interview. 16 subjects started treatment and 1 exited the study for reasons unrelated to treatment. 15 completed. **B)** MADRS scores were obtained weekly and showed a 56% decrease over the course of the study. Of four patients treated at the middle of parcel 46, one out of 4 responded. Of the remaining 11 patients treated at the junction of parcel 46, six out of 11 remitted and 8 out of 11 responded with greater than 50% symptom reduction. **C)** dlPFC-SGACC correlation pre-treatment predicted symptom reduction with all remitters showing close to zero anticorrelation between the two regions. **D)** The area under the ROC curve for response showed a mean area of .88±.11 and sensitivity/specificity of .75 and 1.0 at maximal discrimination. **E)** Percent reduction in peak RFSC significantly correlated with symptom reduction.

The MADRS served as the primary pre-designated outcome measure. Response was defined as a 50% reduction in symptoms. Remission was defined as a final MADRS score ≤10. In addition, HCP-compliant structural and resting-state fMRI (rs-fMRI) scans [6] were acquired pre/post treatment using a Siemens 3T Prisma scanner equipped with 32 channel head coil. These sequences consisted of two T1-weighted MPR vNav scans at 0.8 mm isotropic, matrix = 320×300, slices = 208, TR = 2500 ms, TE = 3.02 ms, flip angle = 8° and two T2-weighted SPC vNav scans at 0.8 mm isotropic, matrix = 320×300, slices = 208, TR = 3200 ms, TE = 564 ms, flip angle = 120°.

The cortical surface of each subject was reconstructed using the function ‘recon-all’ (Freesurfer v6.0.0) with the structural MRI sequences as input. Individual parcellation was obtained by registering the HCPMMP1 atlas labels [6] to each subject’s native space surface using the ‘fsaverage’ surface as an intermediary (https://github.com/tannerjared/HCP-MMP1). The target voxel based on the parcellation results was subsequently annotated on a (native-space) NIFTI file for loading into the navigation software. Rs-fMRI scans were used for pre/post analysis only but not for brain reconstruction.

Eleven individuals received treatment targeting the peak anti-correlation (junction) region. For 3 of these, virtual targeting based on parcel mapping was implemented using individualized 3D-printed scalp headgear [8]. An additional 4 participants received treatment targeting the mid-point of parcel 46. Pre-treatment (MADRS) scores (mean±sem) were 30.9±1.7 and 33.3±2.5 for the 2 groups, respectively. In addition, we evaluated the predictive utility of pre-treatment resting state functional connectivity (rsFC) between DLPFC and sgACC, along with its sensitivity to pre/post change.

Among patients receiving junction-targeted rTMS, there was a 57.8±9.6% reduction in MADRS total score (**Fig. 1B**). Response and remission rates were 72.7% (8/11) and 54.5% (6/11), respectively. Pre-treatment rsFC between parcel 46 and sgACC significantly predicted treatment response, with remitters as a group showing absence of significant anti-correlation between the parcels at baseline (**Fig. 1C**). Moreover, the area under the ROC curve for response showed a mean area of .88±.11 and sensitivity/specificity of .75 and 1.0 at maximal discrimination (**Fig. 1D**). For remission, the corresponding AUC was 1.0. Finally, reductions in rsFC between the peak anti-correlation region and sgACC correlated significantly with treatment response (**Fig. 1E**). Among patients receiving rTMS that targeted the midpoint of parcel 46, we observed a 38.2±19.3% decrease in MADRS, with a response rate of 25% and no remissions, suggesting that small changes in coil position may have a significant clinical effect.

These findings, along with our prior study [7], demonstrate the feasibility of individual, parcel-guided rTMS and suggest response/remission rates similar to that observed using individualized localization of the peak anti-correlated region [9-10]. Notably, in the present study, targeting was performed using structural imaging alone, so the method may be particularly useful in settings in which rs-fMRI scans are unavailable. Further, coupling of the targeting approach with accelerated protocols may further increase response and remission rates [9,10].

In this pilot study, we observed a significant predictive value of baseline rsFC baseline between parcel 46 and sgACC, such that only individuals with reduced (near 0) connectivity at baseline achieved remission. In contrast, individuals with significant anti-correlation at baseline showed lesser improvement. An advantage of the parcel-guided approach is that it permits use of standardized, pre-designated parcels for development of predictive algorithms. Given the small sample size, this finding requires replication in future prospective studies, as well as in post-hoc evaluation of existing pre/post treatment datasets.

## Data Availability

All data produced in the present study are available upon reasonable request to the authors

## Notes

### Competing Interest Statement

The authors declare the following financial interests/personal relationships which may be considered as potential competing interests: JB, DQT, AM, AD, YZV, RM, DCJ report that financial support was provided by National Institutes of Health. DQT, AD, YZV reports a relationship with Soterix Medical Inc that includes: employment. DCJ is inventor on patent #11,779,218 assigned to Columbia University, not bringing in income. Other authors declare that they have no known competing financial interests or personal relationships that could have appeared to influence the work reported in this paper.

### Clinical Trial

NCT04956081

### Author Declarations

The IRBs of Columbia University Medical Center and Western Institutional gave ethical approval for this work

### Funding

- NIH: R44MH126833

